# Preserved recognition of Omicron Spike following COVID-19 mRNA vaccination in pregnancy

**DOI:** 10.1101/2022.01.01.22268615

**Authors:** Yannic Bartsch, Caroline Atyeo, Jaewon Kang, Kathryn J Gray, Andrea G Edlow, Galit Alter

## Abstract

**Background:** SARS-CoV-2 infection is associated with enhanced disease severity in pregnant women. Despite the potential of COVID-19 vaccines to reduce severe disease, vaccine uptake remained relatively low among pregnant women. Just as coordinated messaging from the CDC and leading obstetrics organizations began to increase vaccine confidence in this vulnerable group, the evolution of SARS-CoV-2 variants of concerns (VOC) including the Omicron VOC raised new concerns about vaccine efficacy, given their ability to escape vaccine-induced neutralizing antibodies. Early data point to a milder disease course following omicron VOC infection in vaccinated individuals. Thus, these data suggest that alternate vaccine induced immunity, beyond neutralization, may continue to attenuate omicron disease, such as antibody-Fc-mediated activity. However, whether vaccine induced antibodies raised in pregnancy continue to bind and leverage Fc-receptors remains unclear.

**Methods:** VOC including Omicron receptor binding domain (RBD) or full Spike specific antibody isotype binding titers and FcγR binding were analyzed in pregnant women after the full dose regimen of either Pfizer/BioNtech BNT62b2 (n=10) or Moderna mRNA-1273 (n=10) vaccination using a multiplexing Luminex assay.

**Findings:** Comparable, albeit reduced, isotype recognition was observed to the Omicron Spike and receptor binding domain (RBD) following both vaccines. Yet, despite the near complete loss of Fc-receptor binding to the Omicron RBD, Fc-receptor binding was largely preserved to the Omicron Spike.

**Interpretation:** Reduced binding titer to the Omicron RBD aligns with observed loss of neutralizing activity. Despite the loss of neutralization, preserved Omicron Spike recognition and Fc-receptor binding potentially continues to attenuate disease severity in pregnant women.

**Funding:** NIH and the Bill and Melinda Gates Foundation

## Introduction

While SARS-CoV-2 infection is more likely to cause severe COVID-19 in pregnant individuals, pregnancy was an exclusion criteria from initial vaccine trials, resulting in delayed vaccine roll outs for this medically complex population(*1, 2*). However, following EUA approval of COVID-19 vaccines, eligible pregnant individuals volunteered for vaccine trials and observational studies, providing highly needed data related to safety and immunogenicity to inform vaccine policy. mRNA vaccines proved to be both safe and highly immunogenic when administered throughout pregnancy, giving rise to robust antibody titers that provided critical immunity not only to the mother, but to the infant via transfer of maternal antibodies across the placenta and into breastmilk.

Despite the exciting progress in mRNA vaccine development in pregnancy, the emergence of SARS-CoV-2 variants of concern that can evade neutralizing antibody responses (*3, 4*), including the novel Omicron variant, has led to a global surge in infections (*5*). However, severe disease and death rates have not increased in parallel, pointing to the persistence of alternate vaccine-induced immune responses that may continue to attenuate disease. Among the proposed mechanisms by which vaccine-induced antibodies may still prevent severe disease include, antibody Fc-effector functions, including Fc-mediated opsonophagocytosis and cytotoxicity, which have been linked to survival in severe COVID-19 (*6*) as well as vaccine mediated protection in animal models (*7, 8*).

While pregnant individuals exhibit delayed Fc-effector function maturation after the first dose mRNA vaccine, they elicit a fully functional vaccine-induced Fc-response after the second dose (*9, 10*). However, whether the COVID-19 mRNA vaccines elicit antibody-mediated protection against emerging VOCs, including Omicron, is not well understood. Thus, here we profiled the cross VOC binding titers and Fc-receptor binding profiles after two doses of either the Pfizer/BioNtech BNT162b2 or Moderna mRNA-1273 at peak immunogenicity in a group of fully-vaccinated pregnant individuals.

Despite the decline in IgM, IgA, and IgG binding to the receptor binding domain or full Spike of Omicron compared to other VOCs, Spike-specific Fc*γ*R2a- and Fc*γ*R3a-receptor binding was relatively preserved across both mRNA vaccine platforms for all VOCs, including Omicron. Thus, despite the significant loss of neutralizing antibody responses to Omicron, the preservation of Spike-specific Fc-receptor binding immunity may permit ongoing capture and clearance of the virus even in the absence of strong neutralizing responses to Omicron, thereby providing persistent Fc-mediated protection against severe disease and death.

## Methods

### Study Population

To compare vaccine-induced antibody responses to SARS-CoV-2 VOCs in pregnant individuals, samples from 10 pregnant patients receiving the full two dose regimen BNT162b2 (Pfizer) andr from 10 pregnant patients receiving the full two dose regimen mRNA-1273 (Moderna) were obtained 2-4 weeks after the second dose. All participants were 18 years or older and had an uncomplicated singleton pregnancy. All participants gave informed consent prior to enrollment and this study was approved by the Mass General Brigham Institutional Review Board (IRB; protocol #2020P003538).

### Antigens

Receptor-binding domain antigens for the wildtype (Wuhan), alpha (B.1.1.7), beta (B.1.351), and delta (B.1.617.2) VOCs were obtained from Sino-Biologicals. Omicron RBD was generously provided by Moderna Inc. Stabilized (hexa-pro) Spike of D614G (“wildtype” variant) or VOCs was produced in HEK293 cells.

### IgG subclass, isotype and FcγR binding

Antigen-specific antibody subclass and isotypes, and FcγR binding was analyzed by Luminex multiplexing. The antigens were coupled to magnetic Luminex beads (Luminex Corp, TX, USA) by carbodiimide-NHS ester-coupling with an individual region per antigen. Coupled beads were incubated with different plasma dilutions (1:100 for IgG2, IgG3, IgG4, IgM and IgA1, 1:500 for IgG1 and 1:1,000 for FcγR probing) for 2 hours at room temperature in 384 well plates (Greiner Bio-One, Germany). Unbound antibodies were washed away and subclasses, isotypes were detected with a respective PE-conjugated antibody (anti-human IgG1, IgG2, IgG3, IgG4, IgM or IgA1 all SouthernBiotech, AL, USA) at a 1:100 dilution. For the analysis of FcγR binding PE-Streptavidin (Agilent Technologies, CA, USA) was coupled to recombinant and biotinylated human FcγR2a, FcγR2b, FcγR3a, or FcγR3b protein. Coupled FcγR were used as a secondary probe at a 1:1000 dilution. After 1 h incubation, excessive secondary reagent was washed away and the relative antibody concentration per antigen determined on an IQue analyzer (IntelliCyt).

### Statistical analysis

If not stated otherwise, we assumed non-normal distributions and plots were generated and statistical differences between two groups were calculated in Graph Pad Prism V.8. A Kruskal-Wallis test with a Benjamini-Hochberg post-test correcting for multiple comparisons was used to test for statistical differences between wildtype variant and Omicron titer. Significance was defined as p<0.05. Principal component analysis was performed using the stats (version 3.6.2) package and visualized using the factoextra package (v1.0.7) in R (v.3.6.1) and R Studio (v.1.3).

## Results

### Reduced, but still detectable Omicron-specific isotype immunity following mRNA vaccination

To begin to define whether mRNA vaccines provide protection against distinct VOCs, we first profiled the IgM, IgA, and IgG isotype specific binding capacity of Pfizer/BioNTech BNT162b2 and Moderna mRNA-1273 induced antibodies within pregnant individuals across the wildtype, Alpha, Beta, Delta, and Omicron Receptor Binding Domain (RBD) or whole Spike (S) antigen (Figure 1A and B). Despite the largely preserved Pfizer/BioNTech BNT162b2 and Moderna mRNA-1273 IgM, IgA, and IgG responses to WT, Alpha, Beta, and Delta VOC RBDs, a consistent and significant 16-24-fold (for BNT162b2) and 10-23-fold (for mRNA-1273) loss of IgM, IgA, and IgG binding was noted for vaccine-induced immune responses to the Omicron RBD (Figure 1A). In contrast to the significant decrement in loss of binding against RBD, relatively stable anti-Spike IgM and IgG binding antibodies against all VOCs including Omicron were induced following Pfizer/BioNTech BNT162b2 vaccination in pregnant individuals, while IgA responses were significantly reduced to the Omicron variant (Figure 1B). In contrast to Pfizer/BioNTechBNT162b2, Moderna mRNA-1273 vaccination led to more consistent magnitudes of IgM and IgG responses, and higher IgA responses, in pregnant women across VOCs (Figure1B). However, Moderna mRNA-1273 vaccine responses were significantly lower to Omicron compared to binding levels to the wildtype Spike, across all three isotypes. Yet, despite the more significant loss of Omicron-specific isotype binding in Moderna mRNA-1273-vaccinated pregnant individuals, similar levels of Omicron recognition were observed across both mRNA vaccines due to the overall more uniform or higher titers achieved by the Moderna mRNA-1273 vaccine.

**Figure 1:**
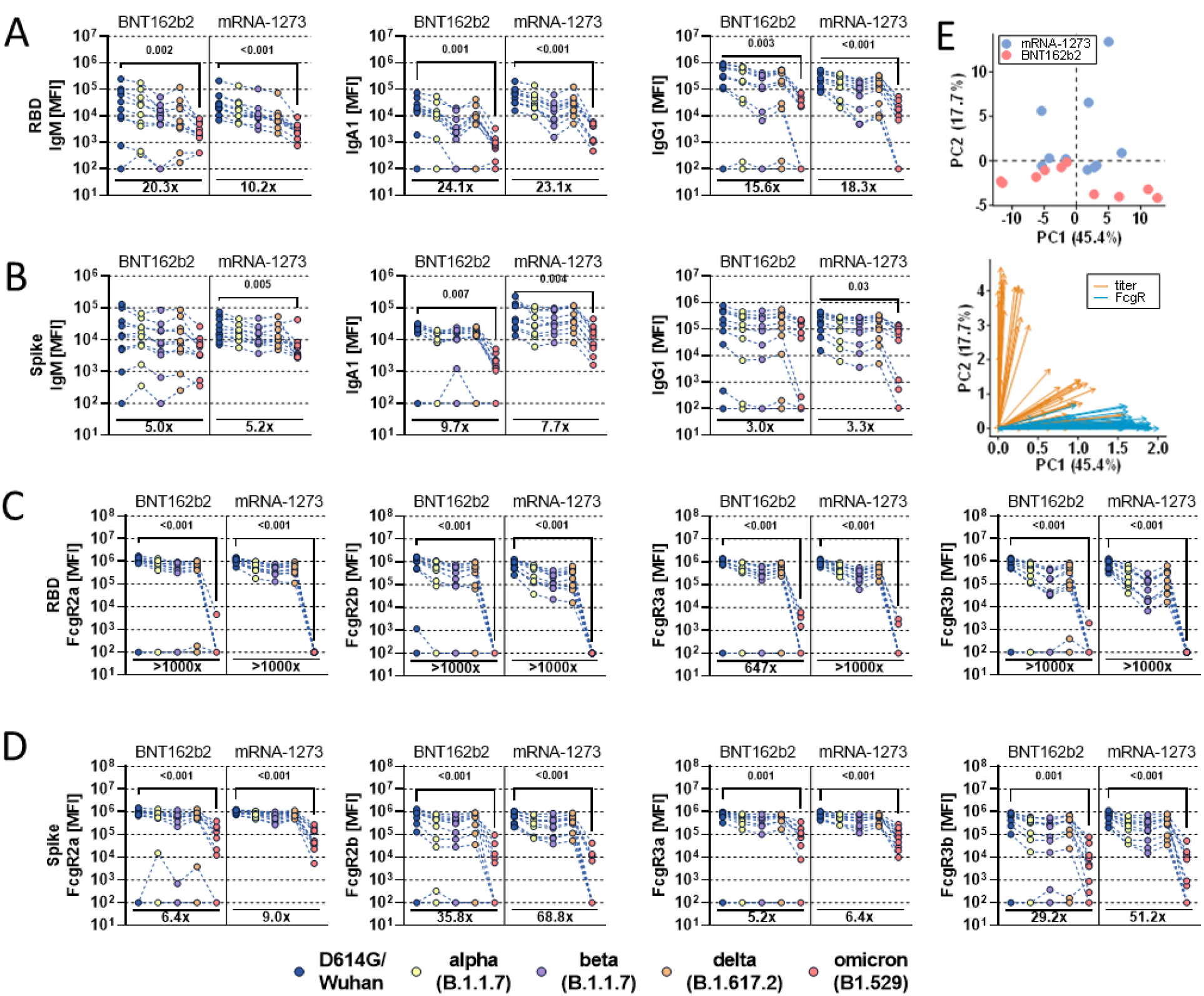
Compromised RBD, but not Spike-specific Omicron recognition in pregnant mRNA vaccinated individuals. Pregnant patients received either the full dose regimen of the BNT162b2(n = 10) or mRNA-1273(n=10). Samples were taken at peak immunogenicity 2-4 weeks after the last dose. IgM, IgA1 and IgG1 binding titers to D614G (WT; blue), Alpha (B1.117; yellow), Beta (B1.351; purple), Delta (B.1.617.2; orange), and Omicron (B1.529; red) variants of concern receptor binding domain (A) or full Spike (B) were measured by Luminex. Binding to FcγR2a, FcγR2b, FcγR3a and FcγR3b of D614G (WT), Alpha (B1.117), Beta (B1.351), Delta (B.1.617.2), and Omicron (B1.529) variant of concern receptor binding domain (C) or full Spike (D) specific antibodies were determined by Luminex. Background corrected data is shown and negative values were set to 100 for graphing purposes in A-D. E) Principal component analysis (PCA) using the complete data set. mRNA-1273 vaccinated individuals are colored blue whereas BNT162b2 recipients are colored in red (upper panel). Loading scores (lower panel) of titer (yellow) or FcγR binding (turquoise) features are plotted along principle component (PC) 1 and 2. The length and direction of the arrow indicates their contribution to the variance explained by the respective PC. A Kruskal-Wallis test with a Benjamini-Hochberg post-test correcting for multiple comparisons was used to test for statistical differences between wildtype variant and Omicron titer. P-values for significant different features are shown above and fold-change reduction of Omicron titer compared to wildtype below each dataset.

### Preserved Spike-, but not RBD-, specific Fc-responses across VOCs

The ability of antibodies to recruit the antiviral activity of the innate immune system, including opsonophagocytic or cytotoxic function, depends on the ability of antibodies to interact with Fc-receptors found on all immune cells (*11*). Among the Fc-receptors, four low affinity Fc-receptors, Fc*γ*R2a, Fc*γ*R2b, Fc*γ*R3a, Fc*γ*R3b, in humans drive IgG mediated activation (*12*). Thus, we next compared the ability of mRNA vaccine induced antibodies to bind VOCs and leverage Fc-receptors. While both BNT162b2 and mRNA-1273 induced RBD-binding antibodies to the wildtype, Alpha, Beta, and Delta VOCs able to bind across all Fc-receptors, Omicron RBD-specific antibodies lost all Fc-receptor binding (Figure 1C). This striking loss of Fc-receptor binding was observed despite of detectable levels of Omicron RBD-specific IgG levels (Figure 1A), highlighting the disconnect between antibody titers and Fc-receptor activation. However, despite this dramatic loss of Omicron RBD-specific Fc-receptor binding, mRNA vaccine-induced Spike-specific antibodies leveraged all Fc-receptors against the wildtype, Alpha, Beta, and Delta variants. Despite losing some Fc-receptor binding capabilities to the Omicron-Spike, both the BNT162b2 and mRNA-1273 vaccine-induced immune responses continued to show detectable binding across all Fc-receptors, with slightly more preserved binding to the activating phagocytic Fc*γ*R2a and cytotoxic Fc*γ*R3a receptors, but a more robust loss of binding to the sole inhibitory Fc*γ*R2b and neutrophil-activating Fc*γ*R3b receptor (Figure 1D). Moreover, when all antibody data were integrated in a multivariate analysis (Figure 1E), BNT162b2 and mRNA-1273 antibody profiles largely overlapped along principal component 1 (which was dominantly explained by FcγR binding) but separated more along PC2 (mainly influenced by titer) highlighting the similar VOC-specific antibody Fc-profiles induced by these vaccines to Omicron and beyond. Collectively, the data highlight a significant loss of Omicron RBD-specific antibody titers and Fc-receptor binding, but the preservation of Omicron Spike-specific antibody binding, that may continue to recognize, clear and control infection, contributing to persistent protection against neutralization-resistant VOCs.

## Discussion

Pregnancy represents an unusual immunological state, in which the maternal immune system must balance tolerance of the fetal graft with protecting the maternal:fetal dyad against foreign pathogens. This delicate balance has been associated with dampened vaccine-induced immune responses (*13*). Specifically, recent deep immunological profiling has pointed not only to differences in the overall magnitude of the vaccine-induced immune response, but also to the kinetics and quality of the evolution of the vaccine induced immune response following COVID-19 mRNA vaccination in pregnancy (*9*). Yet, whether these differences extend to vaccine-induced protection across SARS-CoV-2 variants of concern (VOCs) remains unknown. Moreover, with the emergence of SARS-CoV-2 VOCs, like Omicron, able to evade vaccine-induced neutralizing antibody responses (*3, 4*), defining the ability of antibodies to continue to recognize and leverage the antiviral activity of the immune system may provide critical clues as to vaccine efficacy and need for additional boosters in pregnant individuals. Unfortunately, even predating the emergence of variants of concern that might evade neutralizing antibody responses, vaccine hesitancy has been entrenched and more difficult to combat in the pregnant population (*14-16*), who remain at increased risk for severe COVID-19 (*1, 2*). To date, we are not aware of any studies that have examined efficacy of the mRNA vaccines against new VOCs including Omicron in the pregnant population. Such data are critical to framing and potentially combatting new concerns about vaccine efficacy against Omicron and other VOCs and address the continued importance of vaccination in the pregnant population. While Omicron RBD-specific antibody recognition and Fc-receptor binding were significantly lower compared to other SARS-CoV-2 variants, Spike-specific binding antibodies were able to bind robustly across VOCs, including Omicron, and continued to leverage Fc-receptor binding, pointing to the potential persistence of antibody mediated control of SARS-CoV-2 infection after infection.

Similar to the reported loss of neutralization in the non-pregnant population, here we observed a significant loss of RBD-specific titers and RBD-specific Fc-receptor binding. Conversely, the persistence of Omicron Spike-specific responses, following both the BNT162b2 and mRNA-1273 vaccines, retained the capacity to recruit Fc-receptors, demonstrating the potential for vaccine-induced antibodies to rapidly control and eliminate Omicron infections even in the face of a loss of neutralization. Moreover, the selective preservation of Fc*γ*R2a and Fc*γ*R3a can rapidly leverage phagocytic and cytotoxic immune mechanisms, positioned to rapidly capture and clear opsonized virus upon infection.

While we were unable to comprehensively profile the humoral immune response induced by additional vaccine platforms, or across trimesters of pregnancy, the data presented here provide promising insights on persistent extra-neutralizing properties of mRNA vaccine-induced antibodies that may continue to provide protection against Omicron. Whether boosters can further augment these Fc-receptor recruiting qualities, whether these Fc-functions persist over time, and whether Fc-receptor recruitment alone, in the absence of neutralization, can confer robust protection against Omicron and beyond remains unclear. These initial data may provide critical clues related to the mechanistic correlates of immunity that can guide future vaccine design and boosting in pregnant individuals.

## Data Availability

All data produced in the present work are contained in the manuscript

## Acknowledgments

We thank Nancy Zimmerman, Mark and Lisa Schwartz, an anonymous donor (financial support), Terry and Susan Ragon, and the SAMANA Kay MGH Research Scholars award for their support. We acknowledge support from the Ragon Institute of MGH, MIT, and Harvard, the Massachusetts Consortium on Pathogen Readiness (MassCPR) and the Musk foundation. This study was funded by the NIH (3R37AI080289-11S1, R01AI146785, U19AI42790-01, U19AI135995-02, U19AI42790-01, 1U01CA260476-01, CIVIC75N93019C00052 and The Gates Foundation Global Health Vaccine Accelerator Platform (OPP1146996 and INV-001650). AGE received funding from the NIH (R01HD100022-S2) and March of Dimes Foundation (6-FY-20-223) to support sample collection.

